# Visualizing Decisions and Analytics of Artificial Intelligence based Cancer Diagnosis and Grading of Specimen Digitized Biopsy: Case Study for Prostate Cancer

**DOI:** 10.1101/2022.12.21.22283754

**Authors:** Akarsh Singh, Michael Wan, Lane Harrison, Anne Breggia, Robert Christman, Raimond L. Winslow, Saeed Amal

## Abstract

The rise in Artificial Intelligence (AI) and deep learning research has shown great promise in diagnosing prostate cancer from whole slide image biopsies. Intelligent application interface for diagnosis is a progressive way to communicate AI results in the medical domain for practical use. This paper aims to suggest a way to integrate state-of-the-art deep learning algorithms into a web application for visualizations of decisions and analytics of an AI based algorithms applied on cancer digitized specimen biopsies together with visualizing evidence and explanation of the decision using both image from the biopsy and textual data from Electronic Health Records (EHR). By creating smart visualizations of tissue biopsy images, from magnified regions to augmented sharper images along with image masks that highlight cancerous regions of tissue in addition to intelligent analytics and distribution charts related to cancer prediction, we aim to communicate these easily interpretable results to assist pathologists and concerned medical team to make better decisions for prostate cancer diagnosis as case study.

## 2 INTRODUCTION

Prostate cancer (PCa) is the most common form of cancer among males in the United States. As per the National Institute of Health (NIH) data, globally in 2022, 268,490 new cases of PCa and 34,500 PCa related deaths were reported. AI technologies have shown great promise in the domain of computational healthcare and will prove to be a solid tool for medical examination. Recently deep learning achieved promising results for diagnosing prostate cancer in biopsies [Goldenberg et al. 2019]. Campanella et al. [2018] in their work show that computational pathology and modern deep learning techniques can perform expert-level pathology diagnosis when it comes to cancer detection. However, it is important to make sure the ideas can be communicated and interpreted properly. Imperative healthcare predictions often require integration and visuals to help the healthcare team in better understanding human anatomy [Ooge et al. 2022].

Human involvement in decision-making through visualizations that facilitate interaction with complex algorithms is vital in healthcare. Islam et al. [2019] show that good visualizations elevate communication channels and can be used to discover unknown facts and trends. Data visualization can offer key insights into complicated datasets in ways that are meaningful and intuitive. To visualize their predictions of adverse drug reactions by a logit regression model, Verma et al. [2017] used a snake-like flow diagram. Jönsson et al. [2019] visualized brain regions that significantly differ for people with and without irritable bowel syndrome (IBL), a chronic disease believed to be associated with central brain mechanisms by overlay and highlighting. This inspired us to highlight distinguishing, potential cancerous regions in the biopsies for visualization. Males et al. [2020] Emphasize visual analysis of data from the colon using MRI images and use a zooming technique to enlarge a specific region, compressing the rest. Ko et al. [2017] visualized colon cancer data by creating charts and dashboards, analysing multiple aspects of data under the same bucket. These studies give us great insight for prospective usage of such tools for our application to visualise high resolution biopsies and related meta data. Ghanzouri et al. [2022] facilitate the creation of machine learning (ML) models for diagnosis of peripheral artery disease by wrapping it into an automated tool. Dashboards were created to visualize, risk of peripheral artery disease, relevant factors, patient demographics and guideline recommendations were developed. Nonetheless, on the contrary, uncertainty and randomness in decision making induces mistrust by clinicians, thereby seriously limiting practical usage in actual clinics. Challenges related to standardization, multimodal approaches, and immersive data for medical imaging are well explained by Gillmann et al. [2021]. This article shaped our study and helped us invoke standard practices that are widely acceptable. The science of analytical reasoning supported by the interactive visual interface will intensify the process of bringing AI based technology into critical medical examination. Our goal for this project was to supplement a state-of-the-art deep learning algorithm for prostate cancer classification with a deployed web application and visual summary. The major focus is on the visualizations presented and on quantifying how well are they able to communicate information that will be useful to decision makers (pathologists) to make clinical decisions more accurate and timelier for pathological diagnosis.

## 3 DATA DESCRIPTION

The imaging dataset for this project is based on an open-source deep learning competition hosted on Kaggle named as The Prostate Cancer Grade Assessment Challenge (PANDA) [Bulten et al, 2020]. This competition was the largest of its kind with 12625 whole slide biopsy images (resolution range between 5,000 and 40,000 pixels in both x and y) with masks graded by expert pathologists from two renowned labs. Pathologists characterize tumours into different Gleason growth patterns based on the histological architecture of the tumour tissue. Based on this, biopsy specimens are categorized into one of five groups, International Society of Urologic Pathologists (ISUP) grades. The ISUP grade determined by the sum of the most common primary Gleason pattern (majority) and the highest secondary pattern (minority), as determined by the pathologist on histopathological examination with 3+3 and 5+5 as low and high risk respectively. Figure 1 visualizes the Gleason and ISUP grading process at a higher level. For training, 12625 Whole Slide Images (WSIs) were used. Segmentation masks based on Gleason by hosts model for each data provider were also present for each image in the training set. 940 hidden images were present in the test set for model validation and testing.

**Figure 1:**
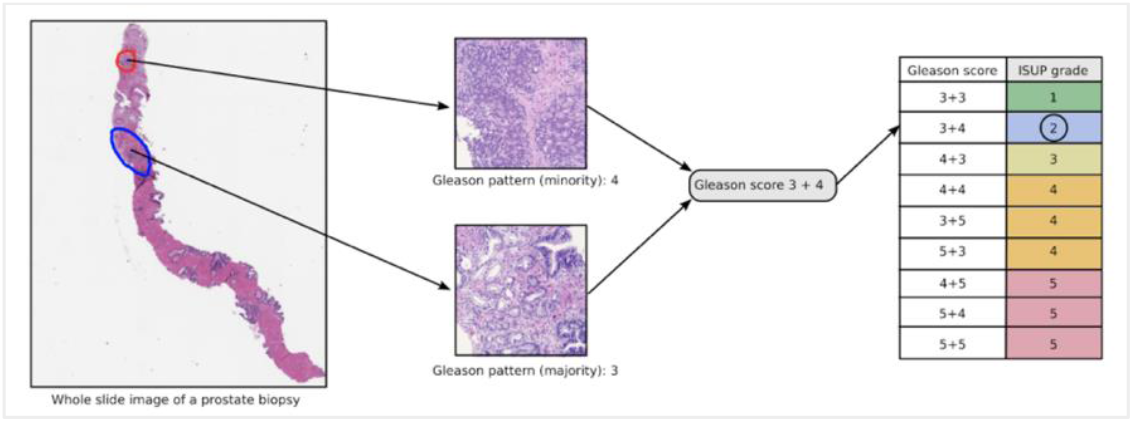
An illustration of the Gleason grading process for an example biopsy containing prostate cancer [Bulten et al, 2020]

## 4 METHODOLOGY

The major workflow progress for this study can be divided into two sections. First, we reproduced the best performing deep learning models and deploying it on an instance (web application). The best performing results came from an ensemble, 5-fold, custom state-of-the-art EfficientNet-B1 [Tan et al. 2019] convolution neural network (CNN) architecture, pre-trained on ImageNet data [Deng et al, 2009]. The models were saved, and a Python script was written to directly use the model pipeline to make predictions on new data. Further, a web application was developed using Flask framework to perform real-time inference. Once a new image was processed and a prediction was made, the respective whole slide image (WSI) was displayed with a predicted grade value (ISUP; 0-5). Second, an approach was taken that emphasizes simple yet powerful visualizations to improve usability and convey the maximum amount of relevant information to the user based on the predicted grade and WSI itself. This aspect involves understanding the data from a visual perspective by performing holistic data analysis and insightful static visualizations. Distribution of the training sample (WSI grades) is shown in Figure 2(a) and 2(b) in the form of bar and pie charts that informs decision makers of what to expect and make statistically driven inferences on the data’s population parameters. We visually explored aspects of the training dataset and delved deep into visualizing prostate (as case study) biopsy WSIs in multiple ways since pathologists analyse them to make diagnosis decisions. Further, we visualized the entire WSI with any option of various resolution scales (corresponding to a down sampling of 1, 4 and 16) as discerning the biopsy slide is how pathologists perform grading and clinical tests.

**Figure 2(a):**
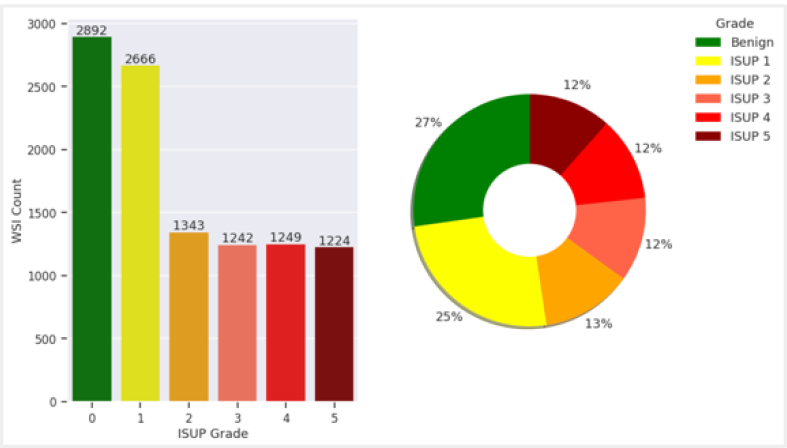
Distribution of Cancer Grade Data

**Figure 2(b):**
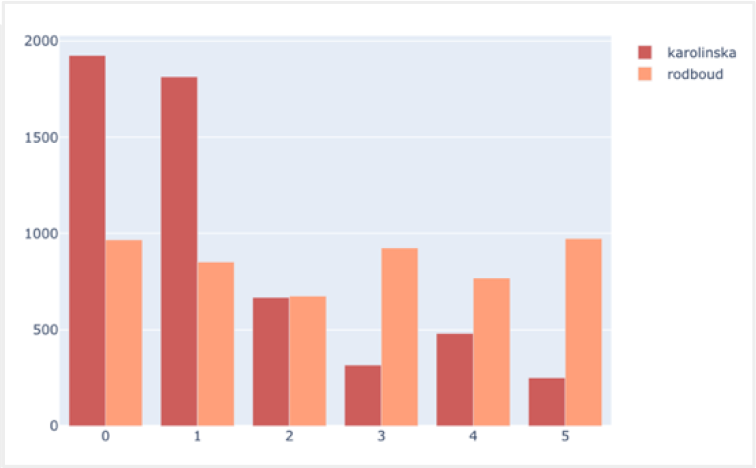
Distribution of Data from Each Hospital

Upon closely observing samples, some have a thick grey boundary along the tissue border (external distortion), sharpened augmentations of the images (Figure 3: Left) were developed to remove it and maximization of cancerous tissue regions. Another reason for this visualization was that due to the grey boundary, the surface of biopsies can often appear hypercellular and can be difficult to interpret [Cahill et al. 2022]. Image masks directly indicate healthy and cancerous areas in the image biopsy. However, since these markups were not present for all images, we plan to integrate the following features in future work. The following (Figure 3: Center and Right) are examples of visualizations that can be created when image masks are available. A side-by-side, highlight visualization was developed (Figure 3: Center) to help find different tissue growth patterns related to cancer that can indicate majority and minority, cancerous subsamples in the WSI. We combined tissue and mask specimens (Figure 3: Right) by overlapping cancerous regions with a light green hue and eliminated insignificant details like surrounding whitespace. This is very important because pathologists grade biopsies for cancer on characteristics like cytoplasm colour (paleness), visible nuclei size, degree of glandular differentiation loss, etc and this overlay helps them identify concerned regions automatically, beforehand. Del Rio et. al [2022] developed similar visualizations from prostate cancer WSIs within an interactive application. To validate the visualizations in terms of usefulness, we intend to share the application with pathologists to understand their needs and obtain invaluable feedback. This will help us better align to their vision and direct us towards a more advanced framework that can provide enhanced visualizations to better support related clinical studies. From initial feedback obtained the visualisations were relevant and their interests lie in exploring quantifiable gains in workflow efficiencies through AI enhanced digital pathology that will be strengthened by constant interpolation and feedback in future work.

**Figure 3:**
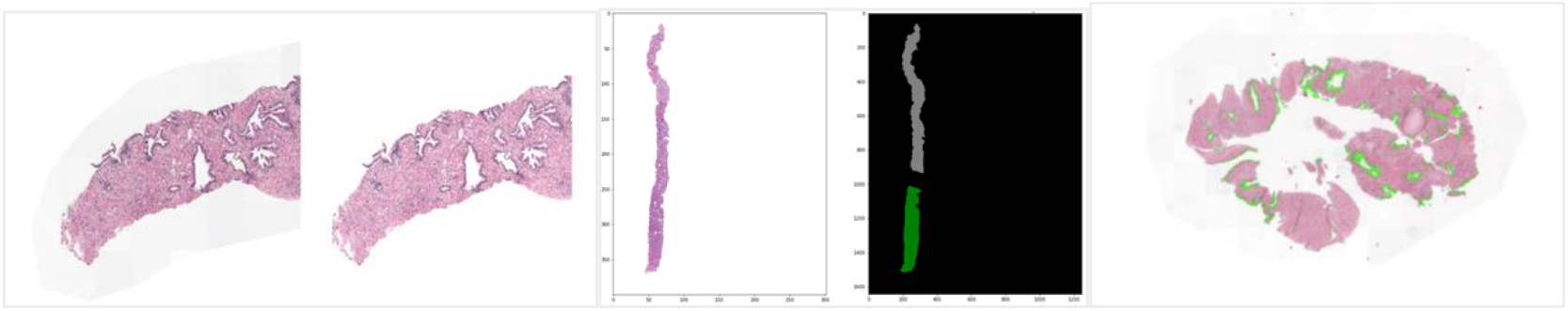
Visualizations of Biopsy Images (Left: Sharpened Image to Eliminate Grey Boundary; Center: Side-by-Side Tissue and Mask Projection; Right: Tissue with Cancerous Overlap)

Finally, donut charts in Figure 4 assist in qualifying prediction results to the decision maker. The left chart outputs SoftMax (activation) probability of each of the 5 deep learning models which constitute our ensemble model, 2 image variations based of pre-processing (variation in image augmentation, resolution, and number of tiles) is considered as input for predictions. This helps in visualizing prediction confidence adding a layer of detail for decision makers to consider how each model is responding in addition to aggregate grade output. The grade corresponding to maximum probability is assumed. All individual model predictions are averaged to give a final prediction. The larger the probability, the higher confidence can be drawn. The right chart takes the final grade prediction and shows the likelihood of it belonging to either of the 2 possible closest grades. For example, here a prediction of 0.21 implies that possibility of the true label being 0 has a 79% chance and 1 has a 21% chance. The prediction of 0.21 is the average prediction from all 5 models. These distributions will help to look for tissue patterns that resemble predicted probability to reach a quicker and more accurate consensus on diagnostic suggestions saving time on grading. All these visualizations combined will steer experts towards the right local tissue regions to look at for diagnosis, save time in the grading process, and determine best recommended diagnosis a patient requires for effective treatment.

**Figure 4:**
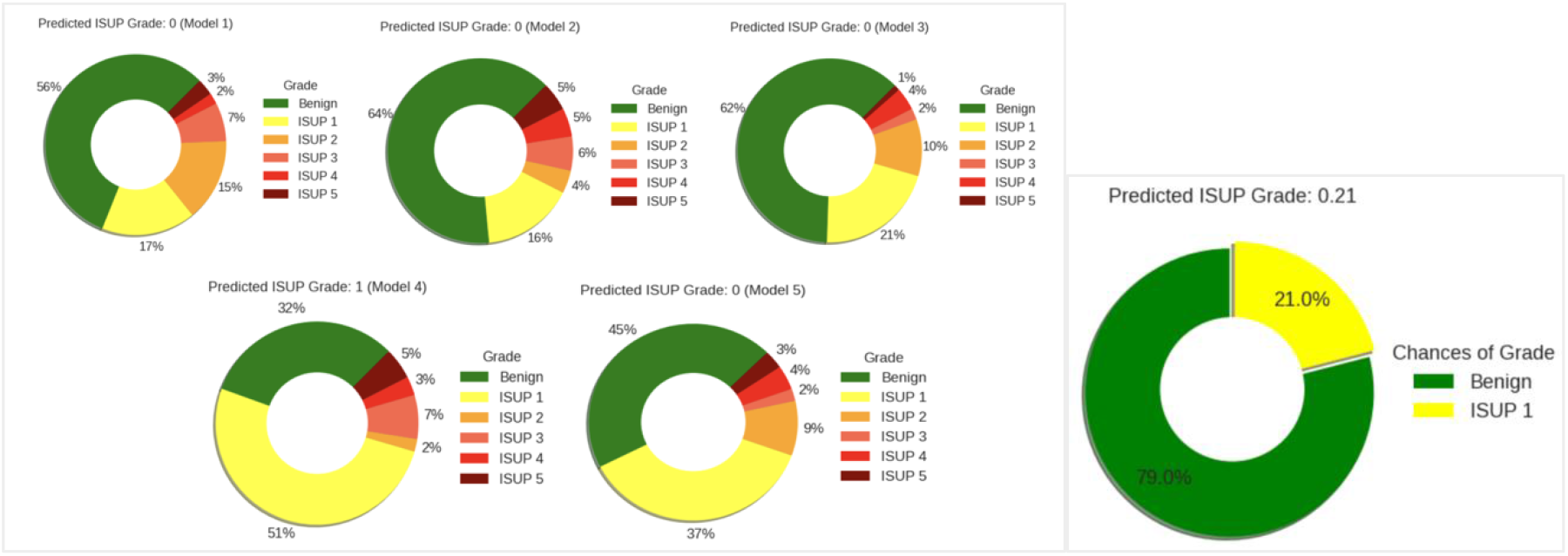
Prediction Output Probability and Ensembled Likelihood Distribution

## 5 RESULTS AND FUTURE SCOPE

Projected visualizations are powerful and well capable of highlighting cancerous tumour regions in available biopsies, image augmentations, sharpness, and contrast changes help improve resolution, especially where specific areas of tissues are concerned. Multiple levels of AI-assisted WSI visualizations spanning from concentrated resolutions of local tissue properties to the entire biopsy with cancerous overlap serve as a great tool to assist in accurate diagnosis of prostate cancer and reducing pathological workload. The technology capabilities in tandem with the web application and powerful visualization set the stage for a demonstratable product to be used in practice. Moving forward, we aim to test the application with collaborators, clinics and obtain further feedback on the interface, application smoothness, interpretability, and communication strength (from experienced pathologists). This can be quantified by working feedback surveys and ratings in critical aspects. In later stages, we also plan on obtaining more data to fine tune our deep learning models to obtain enhanced metric scores for visualizations with further enhanced precision. Moreover, by establishing trust in practice, we can work on generalizing all aspects of the application i.e., backend machine learning code, security, visualizations and user interface in varied clinical settings.

## Data Availability

All data produced in the present work are contained in the manuscript

